# Advantages of DDG-certified hospitals for hospitalized patients with diabetes - A nationwide DRG analysis in Germany

**DOI:** 10.1101/2025.04.07.25325368

**Authors:** Marie Auzanneau, Andreas Fritsche, Alexander J. Eckert, Esther Seidel-Jacobs, Martin Heni, Stefanie Lanzinger

**Affiliations:** Institute of Epidemiology and Medical Biometry, Ulm University, Ulm, German; German Center for Diabetes Research (DZD), Neuherberg, Germany; Department of Internal Medicine IV, University Hospital Tübingen, Germany; Institute of Diabetes Research and Metabolic Diseases (IDM) of the Helmholtz Center Munich at the University of Tübingen, Tübingen, Germany; Institute for Biometrics and Epidemiology, German Diabetes Center (DDZ), Leibniz Center for Diabetes Research at Heinrich-Heine-University Düsseldorf, Düsseldorf, Germany; Institute for Clinical Chemistry and Pathobiochemistry, Department for Diagnostic Laboratory Medicine, University Hospital Tübingen, Tübingen, Germany; Division of Endocrinology and Diabetology, Department of Internal Medicine 1, University Hospital Ulm, Ulm, Germany

**Author notes:** <u>Corresponding Author:</u>* Marie Auzanneau, MPH, PhD, Institute of Epidemiology and Medical Biometry, Ulm University, Albert-Einstein-Allee 41, D-89081 Ulm, Germany, Tel.: +49/731/5025483, Fax: +49/ 731/5025309.

## Abstract

**Aim:** Hospital certifications by the German Diabetes Association (DDG) are intended to ensure high-quality diabetological care. We aimed to compare Germany-wide characteristics and outcomes of hospitalized adults with diabetes in diabetes certified vs. non-diabetes certified hospitals.

**Methods:** We analyzed in the German Diagnosis Related Groups (DRG) statistics for 2021-2023 all inpatient cases aged ≥ 20 years with and without diabetes (all types) as a main or secondary diagnosis based on ICD-10 codes. Using Wilcoxon and Chi^2^-tests, characteristics of inpatient cases with diabetes were compared between diabetes certified hospitals (n=300) and non-diabetes certified hospitals (n=1,103).

**Results:** Of 43.4 million total inpatient cases in the period from 2021 to 2023, 8.1 million (18.5%) had a documented diabetes diagnosis (94% of the cases as secondary diagnosis). In certified hospitals, 19.0% of inpatient cases had diabetes compared to 18.3% in non-certified hospitals. Although inpatient cases with diabetes had more hospital-acquired and procedure-related complications (17.2% vs. 16.7%, p<0.001), as well as higher rates of hypoglycemia (3.2% vs. 2.1%, p<0.001) and metabolic disorder (diabetic ketoacidosis: 0,5% vs. 0,4%; acute metabolic disorder with multiple complications: 2.3% vs. 1.0%, p<0.001) in certified hospitals vs. non-certified hospitals, in-hospital mortality for all diabetes cases was similar (4.5% in both). However, among those with diabetes as the main diagnosis, in-hospital mortality was significantly lower in diabetes certified hospitals (type 1 diabetes: 0.6% vs. 1.1% and type 2 diabetes: 2.3% vs. 2.9%, both p<0.001), despite a higher complication burden.

**Conclusions:** Hospitalized people with diabetes as the main diagnosis had lower in-hospital mortality in diabetes-certified hospitals, even though they had a higher disease burden. These findings suggest that expanded use of DDG-certified departments, including consultation services for patients whose diabetes is a secondary diagnosis, may further improve clinical outcomes.

## Introduction

Diabetes is highly prevalent in Germany, with more than 9 million people living with diagnosed condition (1,2), and at least 2 million more estimated to have undiagnosed diabetes (1). According to the International Diabetes Federation (IDF), Europe had the second highest number of diabetes-related deaths in 2021 (approximately 1.1 million) after the Western Pacific region (3).

Although diabetes is typically managed in an outpatient setting, one in five hospitalized people in Germany has a diabetes diagnosis (4). Moreover, up to 40% of hospitalized people in internal medicine, surgery and neurology departments of maximum care hospitals have diabetes (5), underscoring the importance of effective inpatient diabetes management.

In Germany as in many other countries, diabetes is associated with a high economic and health burden for patients and represents a major cost factor for the healthcare system (3,6,7). It is associated with need for surgery at a younger age, more frequent complications, prolonged hospital stays, and higher in-hospital mortality (4,8). By 2040, an estimated 11 to 12 million people in Germany will be living with diabetes (2), posing serious challenges for the healthcare system. However, improved diabetes treatment can lead to significant health benefits and reduce overall healthcare costs (6). In several countries, including Denmark (9), Belgium (10) or Australia (11), implementation of diabetes-specific hospital accreditations has led to improvements in the diabetes care.

In Germany, the German Diabetes Association (DDG) confers certifications for hospitals that can demonstrate evidence-based, guideline-compliant diabetes treatment and have appropriately qualified medical personnel (12). The aim of this analysis was to compare the characteristics and outcomes of hospitalized people with diabetes between diabetes-certified hospitals (DCH) versus non-diabetes-certified hospitals (NDCH) for the years 2021 to 2023. These results should help to design future prospective studies and help optimizing inpatient diabetes care across Germany.

## Methods

### Data source

All inpatient cases aged ≥ 20 years with and without diabetes as a main or secondary diagnosis were analyzed based on ICD-10 codes in the nationwide diagnosis-related groups (DRG) statistics for 2021-2023, collected by the German Federal Statistical Office and Statistical Offices of the Federal States. A diagnosis of diabetes included type 1 diabetes or T1D (E10), type 2 diabetes or T2D (E11), other specified diabetes mellitus including pancreatic diabetes (diabetes resulting from diseases of the exocrine pancreas) (E13), rare types of diabetes (E12 or E14), and gestational diabetes (O24).

### Identification of the DCH

We used the list of hospitals certified by the DDG (n=867 as of 22.11.2023, https://www.ddg.info/fileadmin/user_upload/20231122_Zert.Einrichtungen.xlsx) to select all hospitals that had received at least one of the DDG certificates (“Diabetes Exzellenzzentrum DDG”, “Diabeteszentrum DDG”, “Klinik mit Diabetes im Blick DDG” oder “Zertifizierte Fußbehandlungseinrichtung DDG”, (12)) at any time during the period 2021-2023 (n=425). We selected only hospital providing inpatient care, excluding outpatient centers as well as inpatient rehabilitation centers. We complemented the list with the identification code of each institution (“IK-Nummer”, available from the Quality report of the hospitals: https://www.g-ba.de/themen/qualitaetssicherung/datenerhebung-zur-qualitaetssicherung/datenerhebung-qualitaetsbericht/) and grouped different names with the same code as one institution. We finally obtained a list of 311 DCH with different identification codes. These codes were used to categorize the 1,415 hospitals from the DRG statistics for 2021-2023 in DCH and NDCH. We only included hospitals in the analysis that coded at least one diabetes case in the period 2021-2023 (n=1,403 hospitals, of which 300 DCH and 1,103 NDCH).

### Characteristics and outcomes

We compared the following parameters for inpatient cases with diabetes between DCH and NDCH: age, sex, length of hospital stay (days), in-hospital mortality (documentation of “death” as the reason for discharge from hospital), and searched in the main or secondary diagnoses ICD-10 codes for obesity (E66), hospital-acquired and procedure-related complications (table 4, list of ICD-codes), hypoglycemia (E10-14.6), diabetic ketoacidosis, DKA (E10-14.1), and acute metabolic disorders with multiple complications (E10-14.73).

The results were compared using Wilcoxon-tests for continuous variables or Chi2-tests for binary variables. P-values were adjusted for multiple testing according to the Holm-Bonferroni method. The level of significance was set at 0.01 (two-sided). All analyses were performed with SAS version 9.4 (build TS1M8; SAS Institute, Inc., Cary, NC) on a windows server mainframe.

## Results

Overall, the nationwide DRG statistics recorded 43.4 million inpatient cases with or without diabetes from 2021 to 2023 (on average 14.5 million per year). Of these inpatient cases, 38% (n=16,688,051) took place in the 21% of hospitals (n=300) with DDG certification. Consequently, one DCH handled more inpatient cases (with or without diabetes) than one NDCH (on average, 18,542 cases per hospital per year vs. 8,085 cases per hospital per year).

During these three years, a main or secondary diagnosis of diabetes (all types) has been coded in 18.5% of all inpatient cases (n= 8,055,686 inpatient cases with diabetes, including 2,045 cases with T1D and T2D as double secondary diagnoses).

The absolute frequency of inpatient cases with diabetes was on average more than twice as high at a certified hospital than at a non-certified hospital (DCH: n= 3,519 per hospital per year vs. NDCH: n=1,477 per hospital per year). This corresponded also to a higher proportion of inpatient cases with diabetes in the certified hospitals (DCH: 19.0% vs. NDCH: 18.3%, p<0.001). Among diabetes cases treated in DCH compared to the NDCH (table 1), patients were slightly younger (71 vs. 72 years old, p<0.001) and more often male (56% vs. 55%, p<0.001). They also had higher rates of hospital-acquired and procedure-related complications (17.2% vs. 16.7%, p<0.001), hypoglycemia (3.2% vs. 2.1%, p<0.001), and other acute metabolic complications (DKA: 0.5% vs. 0.4% and acute metabolic disorder with multiple complications: 2.3% vs. 1.0%, p<0.001). However, the length of stay and the in-hospital mortality was similar between DCH and NDCH (4.5% vs 4.5%, p=0.061).

**Table 1:**
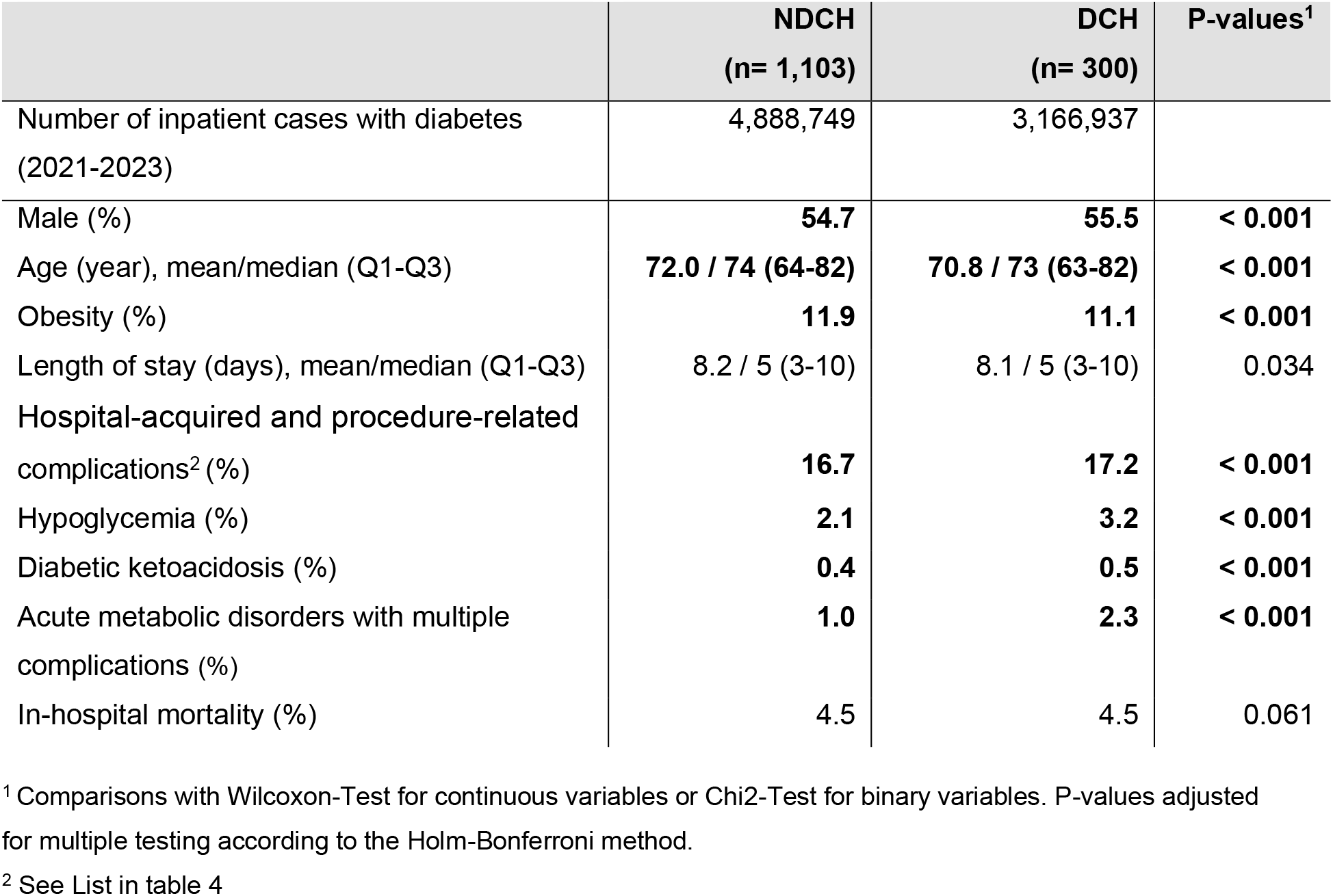
Comparison of inpatient cases with diabetes as main or secondary diagnosis in non-diabetes-certified hospitals (NDCH) and diabetes-certified hospitals (DCH)

|  | NDCH<br>(n= 1,103) | DCH<br>(n= 300) | P-values<sup>1</sup> |
| --- | --- | --- | --- |
| Number of inpatient cases with diabetes (2021-2023) | 4,888,749 | 3,166,937 |  |
| Male (%) | 54.7 | 55.5 | < 0.001 |
| Age (year), mean/median (Q1-Q3) | 72.0 / 74 (64-82) | 70.8 / 73 (63-82) | < 0.001 |
| Obesity (%) | 11.9 | 11.1 | < 0.001 |
| Length of stay (days), mean/median (Q1-Q3) | 8.2 / 5 (3-10) | 8.1 / 5 (3-10) | 0.034 |
| Hospital-acquired and procedure-related complications<sup>2</sup> (%) | 16.7 | 17.2 | < 0.001 |
| Hypoglycemia (%) | 2.1 | 3.2 | < 0.001 |
| Diabetic ketoacidosis (%) | 0.4 | 0.5 | < 0.001 |
| Acute metabolic disorders with multiple complications (%) | 1.0 | 2.3 | < 0.001 |
| In-hospital mortality (%) | 4.5 | 4.5 | 0.061 |

<sup>1</sup> Comparisons with Wilcoxon-Test for continuous variables or Chi2-Test for binary variables. P-values adjusted for multiple testing according to the Holm-Bonferroni method.
<sup>2</sup> See List in table 4

### Leading Reason for hospital admission

In both DCH and in NDCH, cardiovascular disease was the most frequent reason for admission in cases with diabetes. Six (NDCH) to seven (DCH) of the ten most frequent main diagnoses belonged to the ICD group of the “disease of the cardiovascular system (I00-I99)” *(table 2)*.

**Table 2:**
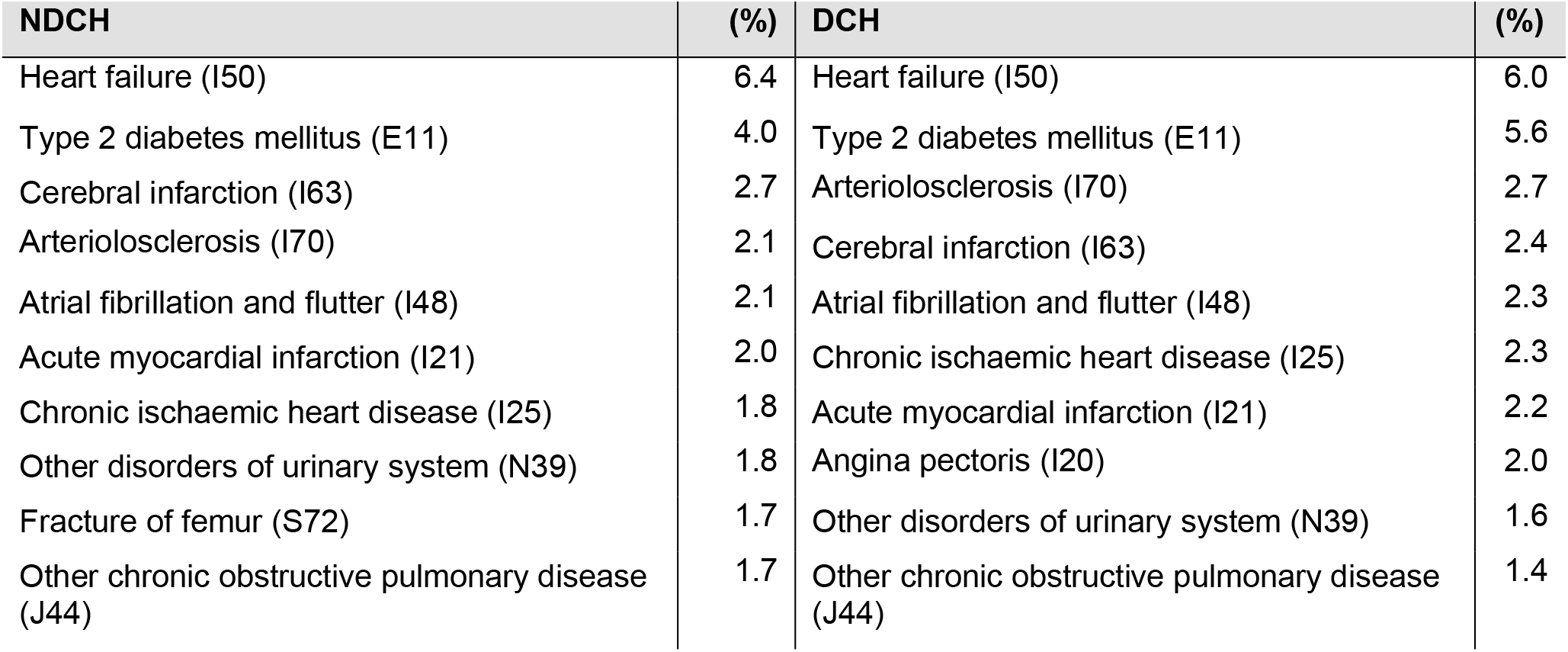
Most frequent reasons for admission (main diagnoses) for inpatient cases with diabetes in non-diabetes-certified hospitals (NDCH) and diabetes-certified hospitals (DCH)

Diabetes itself was infrequently documented as the reason for admission (main diagnosis): only 0.2% of all inpatient cases had a T1D as main diagnosis (n= 71,875) and 0.9% a T2D (n= 374,273) (*table 3)*.

**Table 3a:**
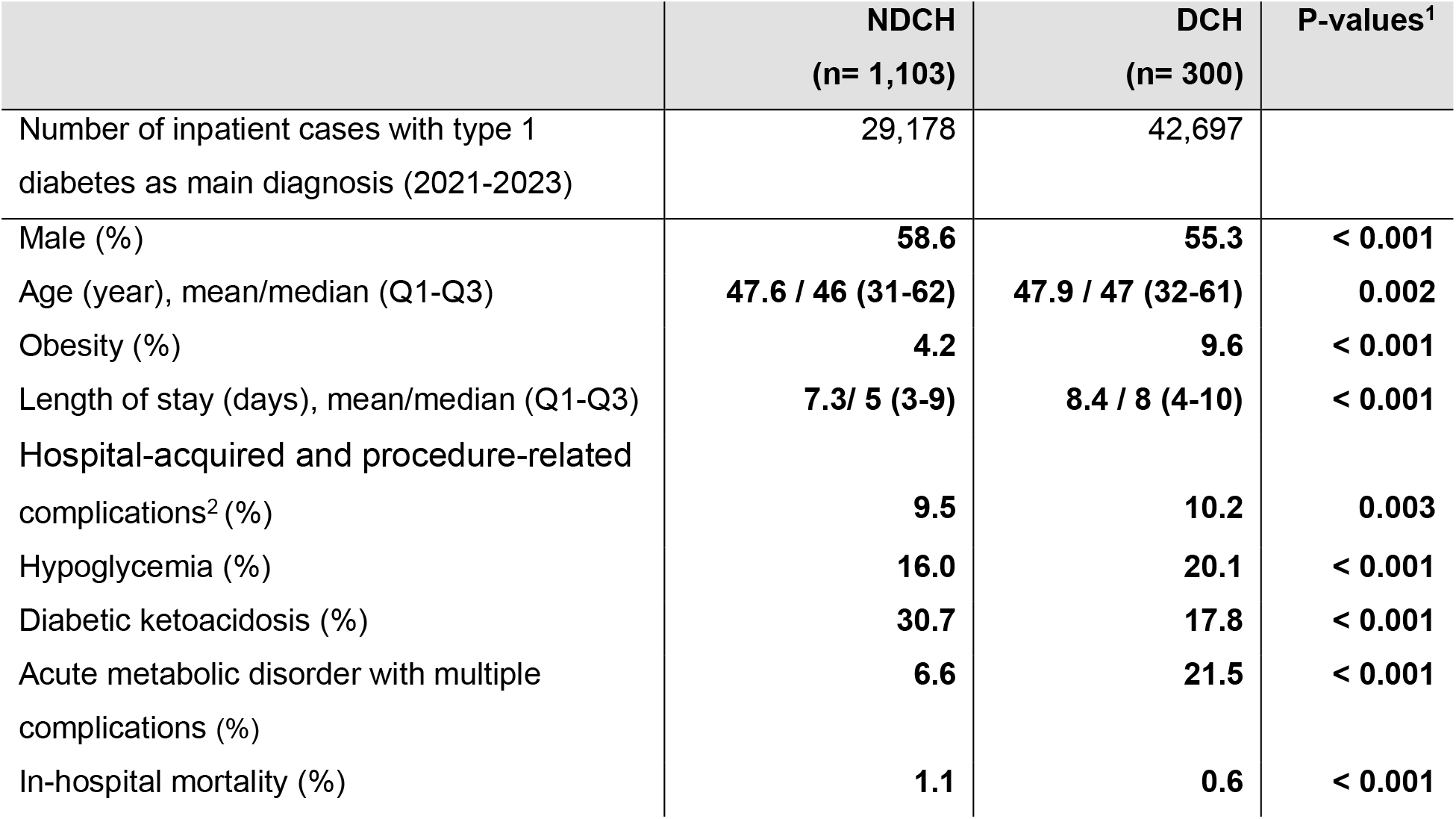
Comparison of inpatient cases with type 1 diabetes as main diagnosis in non-diabetes-certified hospitals (NDCH) and diabetes-certified hospitals (DCH)

|  | **NDCH**<br>**(n= 1,103)** | **DCH**<br>**(n= 300)** | **P-values<sup>1</sup>** |
| --- | --- | --- | --- |
| Number of inpatient cases with type 1 diabetes as main diagnosis (2021-2023) | 29,178 | 42,697 |  |
| Male (%) | **58.6** | **55.3** | **< 0.001** |
| Age (year), mean/median (Q1-Q3) | **47.6 / 46 (31-62)** | **47.9 / 47 (32-61)** | **0.002** |
| Obesity (%) | **4.2** | **9.6** | **< 0.001** |
| Length of stay (days), mean/median (Q1-Q3) | **7.3/ 5 (3-9)** | **8.4 / 8 (4-10)** | **< 0.001** |
| Hospital-acquired and procedure-related complications<sup>2</sup> (%) | **9.5** | **10.2** | **0.003** |
| Hypoglycemia (%) | **16.0** | **20.1** | **< 0.001** |
| Diabetic ketoacidosis (%) | **30.7** | **17.8** | **< 0.001** |
| Acute metabolic disorder with multiple complications (%) | **6.6** | **21.5** | **< 0.001** |
| In-hospital mortality (%) | **1.1** | **0.6** | **< 0.001** |

**Table 3b:** Comparison of inpatient cases with type 1 diabetes as main diagnosis in non-diabetes-certified hospitals (NDCH) and diabetes-certified hospitals (DCH)

|  | **NDCH**<br>**(n= 1,103)** | **DCH**<br>**(n= 300)** | **P-values<sup>1</sup>** |
| --- | --- | --- | --- |
| Number of inpatient cases with type 2 diabetes as main diagnosis (2021-2023) | 196,686 | 177,587 |  |
| Male (%) | **62.8** | **64.7** | **< 0.001** |
| Age (year), mean/median (Q1-Q3) | **71.2 / 73 (62-82)** | **69.2 / 71 (60-80)** | **< 0.001** |
| Obesity (%) | **12.1** | **18.9** | **< 0.001** |
| Length of stay (days), mean/median (Q1-Q3) | **9.9 / 7 (4-12)** | **10.9 / 8 (5-13)** | **< 0.001** |
| Hospital-acquired and procedure-related complications<sup>2</sup> (%) | **16.6** | **21.0** | **< 0.001** |
| Hypoglycemia (%) | **14.6** | **13.2** | **< 0.001** |
| Diabetic ketoacidosis (%) | **3.2** | **2.5** | **< 0.001** |
| Acute metabolic disorder with multiple complications (%) | **3.9** | **10.5** | **< 0.001** |
| In-hospital mortality (%) | **2.9** | **2.3** | **< 0.001** |

<sup>1</sup> Comparisons with Wilcoxon-Test for continuous variables or Chi2-Test for binary variables. P-values adjusted for multiple testing according to the Holm-Bonferroni method.
<sup>2</sup> See list in table 4

Among all inpatient cases with diabetes, T1D as a main diagnosis was twice as often documented in DCH (1.3%) than in NDCH (0.6%). Hence, 59.4% of all inpatient cases with T1D as a main diagnosis were treated in DCH. These patients treated in DCH vs. NDCH were slightly older (47.9 vs. 47.6 years old, p=0.002) and less frequently male (55 vs. 59%, p<0.001). In addition, they had more episodes of hypoglycemia (20.1% vs. 16.0%) and acute metabolic disorders with multiple complications (21.5% vs. 6.6%, both p<0.001). However, DKA was less common (17.8% vs. 30.7%, p<0.001).

Among all inpatient diabetes cases, T2D as a main diagnosis was also more often documented in DCH (5.6%) than in NDCH (4.0%). Thus, 47.4% of all inpatient cases with T2D as a main diagnosis were treated in DCH. These inpatients treated in DCH vs. NDCH were younger (69.2 vs. 71.2 years old, p<0.001) and more often male (65 vs. 63%, p<0.001). In addition, they had more hospital-acquired and procedure-related complications (21.0% vs. 16.6%), and acute metabolic disorders with multiple complications (10.5% vs. 3.9%; both p<0.001).

### Obesity, length of stay, and in-hospital mortality

Inpatients with T1D and T2D as the main diagnosis in DCH vs. NDCH presented more frequently with obesity (T1D: 9.6% vs. 4.2%; T2D: 18.9% vs. 12.1%; both p<0.001). They had a longer hospitalization (by about one day in T1D and T2D, both comparisons: p<0.001). Despite this, in-hospital mortality was lower in DCH for both T1D (0.6% vs. 1.1%) and T2D: (2.3% vs. 2.9%; both p<0.001).

## Discussion

This nationwide analysis of the mandatory DRG statistics confirms for the years from 2021 to 2023 the result of our previous publication revealing that almost one in five hospitalized patients in Germany carries a documented diagnosis of diabetes mellitus (4). Given the high number of unrecognized diabetes (1) and the potential underreporting of the secondary diagnosis of diabetes in the DRG statistics, the true prevalence of hospitalized patients with diabetes is probably even higher (5).

Our findings indicate that DCH handled more overall inpatient cases per hospital than the NDCH and thus represent larger hospitals. In children and adolescents with diabetes, it has already been shown that the size of the hospital influences the quality of outpatient care: acute complications were less frequent in larger centers than in smaller centers (13).

Regarding for adults with diabetes, it has also been reported that larger hospitals use significantly less insulin monotherapy than smaller hospitals, which indicates differences in care by center size (14).

On average, one DCH had more than twice as many hospitalizations with diabetes per year than one NDCH. Not only the absolute frequency per hospital but also the proportion of inpatient cases with diabetes were higher in DCH compared to NDCH. In addition, DCH documented diabetes (particularly T1D) more frequently as the main diagnosis. This could reflect both greater disease severity at admission and a heightened awareness of diabetes as a primary reason for hospitalization. Nevertheless, even if one DCH had more inpatient cases with diabetes on average than a NDCH, 61% of all hospitalized diabetes patients in Germany still receive care in non-certified centers.

Consistent with prior research (4), cardiovascular disease was the leading reason for admission among people with diabetes, both in DCH and NDCH. Indeed, diabetes is associated with a 2 to 4-fold increased risk of cardiovascular disease, such as coronary heart disease, cerebrovascular complications, heart failure or peripheral arterial disease (15).

Notably, obesity appears to be undercoded (appearing in only 11-12% of all diabetes cases), although 92.3% of inpatient cases with diabetes had T2D. The prevalence of obesity in people with T2D (45 to 79 years) in Germany is estimated at 54% (16). This likely undercoding in documentation practices in cases with diabetes may result from the coding of mainly cases of extreme obesity (17). Interestingly, obesity was coded more frequently in people with a main diagnosis of diabetes, especially in the DCH. It is possible that more obese people with diabetes and especially more people with more severe obesity are hospitalized in DCH. It is also possible that the quality of documentation of a secondary diagnosis of obesity is better in DCH.

Our analyses reveal that patients in DCH generally presented with more complications, hypoglycemia, and acute metabolic disorders, suggesting a higher severity of illness. Even those with T1D or T2D as the main diagnosis were more likely to have obesity, complications, and acute metabolic disorders with multiple complications in DCH. Nevertheless, we observed no difference in in-hospital mortality between DCH and NDCH when considering all diabetes cases (main and secondary diagnosis). Of note, in the subset of patients whose main diagnosis was diabetes, in-hospital mortality was even lower in DCH, for both T1D and T2D, and the proportion of DKA was almost halved in those with a main diagnosis of T1D. This suggests that specialized expertise available in diabetes-certified units, including diabetology departments and consultation services, may improve outcomes for patients most actively managed for diabetes.

### Strengths and limitations

A major strength of this study is the use of mandatory nationwide DRG data, which captures almost all acute-care hospital admissions in Germany (except for psychiatric and psychosomatic hospitals and rehabilitation facilities). Including both main and secondary diagnoses of diabetes, this study describes the real prevalence of documented diabetes in hospitals. Nonetheless, the DRG statistics refer to hospital admissions (“inpatient cases”) and do not provide any information at patient level. As the DRG system was designed primarily for billing rather than research, the quality of the coding is not guaranteed. However, systematic overcoding of diabetes for billing reasons is unlikely because it is currently hardly relevant to revenue in the DRG system. However, undercoding of diabetes and other comorbidities as secondary diagnoses, which are hardly relevant for billing purposes, cannot be ruled out.

These data provide indications for the comparison of hospitalized people with diabetes between the DCH and NDCH. However, it is difficult to draw conclusions about differences in quality of care depending on certification for several reasons. First, our results suggest that more people with more severe conditions are hospitalized in DCH, which are larger hospitals overall, and probably more often maximum care hospitals. Unfortunately, the DRG statistics does not provide this information. This potential selection bias prevents us from comparing treatment outcomes between DCH and NDCH directly.

A further limitation is that the identification of the DCH by the institution code of the hospitals does not make it possible to differentiate between the different types of DDG certificates, as different departments of a hospital can have different certificates under a single institution code. It is also not possible to draw conclusions about which specialist departments within a hospital are actually certified for diabetes based on the institution code. The vast majority of people with diabetes who were treated as inpatients were admitted for other main diagnoses. In the DCH, too, the majority of diabetes patients were therefore probably not treated in the diabetes departments that are certified, but in other departments. However, it should be assumed, at least in part, that people with diabetes admitted to DCH as inpatients were in most departments also treated for their diabetes by a diabetology-certified consultation service (“diabetes unit”).

## Conclusion

These findings suggest that treatment in DCH particularly benefits patients admitted primarily with diabetes who even experienced lower in-hospital mortality, likely due to broad diabetes expertise. However, since most hospitalized people with diabetes are admitted for other conditions, extending specialized care to secondary-diagnosis cases could further help reduce complications and improve outcomes. Implementing hospital-wide diabetes consultation services, expanding structured diabetes teams, and establishing a hospital-wide diabetes unit could align inpatient care with DDG standards, potentially improving outcomes hospital-wide.

## Data Availability

The German Diagnosis Related Groups (DRG) statistics for 2021-2023 (Source: Research data centre (RDC) of the Federal Statistical Office and Statistical Offices of the Federal States, DOI [10.21242/23141.2021.00.00.6.1.0; 10.21242/23141.2022.00.00.6.1.0; 10.21242/23141.2023.00.00.6.1.0]), is available against fee via controlled remote analysis through the RDC of the Federal Statistical Office.

## Supplemental data

**Table 4:** ICD-10 codes and diagnoses that were combined as “Hospital-acquired and procedure-related complications”.

## Data source

Research data centre (RDC) of the Federal Statistical Office and Statistical Offices of the Federal States, DOI [10.21242/23141.2021.00.00.6.1.0; 10.21242/23141.2022.00.00.6.1.0; 10.21242/23141.2023.00.00.6.1.0], own calculations.

## Acknowledgements

Special thanks to Andreas Hungele and Ramona Ranz (clinical data managers, Ulm University) for their support and the development of the DPV documentation software. We express our sincere gratitude to Prof. Reinhard Holl, the initiator of the DPV registry, for his achievements.

