## Supplemental Table 4 for "Advantages of DDG-certified hospitals for hospitalized patients with diabetes - A nationwide DRG analysis in Germany"

**Supplemental table 4. ICD-10 codes and diagnoses that were combined as "Hospital-acquired and procedure-related complications".**

| ICD-10 Code | Diagnoses |
|---|---|
| **E89.0** | Postprocedural hypothyroidism |
| **E89.1** | Postprocedural hypoinsulinaemia |
| **E89.2** | Postprocedural hypoparathyroidism |
| **E89.3** | Postprocedural hypopituitarism |
| **E89.4** | Postprocedural ovarian failure |
| **E89.5** | Postprocedural testicular hypofunction |
| **E89.6** | Postprocedural adrenocortical(-medullary) hypofunction |
| **E89.8** | Other postprocedural endocrine and metabolic disorders |
| **E89.9** | Postprocedural endocrine and metabolic disorder, unspecified |
| **G97.0** | Cerebrospinal fluid leak from spinal puncture |
| **G97.1** | Other reaction to spinal and lumbar puncture |
| **G97.2** | Intracranial hypotension following ventricular shunting |
| **G97.8** | Other postprocedural disorders of nervous system |
| **G97.9** | Postprocedural disorder of nervous system, unspecified |
| **H59.0** | Keratopathy (bullous aphakic) following cataract surgery |
| **H59.8** | Other postprocedural disorders of eye and adnexa |
| **H59.9** | Postprocedural disorder of eye and adnexa, unspecified |
| **H95.0** | Recurrent cholesteatoma of postmastoidectomy cavity |
| **H95.1** | Other disorders following mastoidectomy |
| **H95.8** | Other postprocedural disorders of ear and mastoid process |
| **H95.9** | Postprocedural disorder of ear and mastoid process, unspecified |
| **I97.0** | Postcardiotomy syndrome |
| **I97.1** | Other functional disturbances following cardiac surgery |
| **I97.2** | Postmastectomy lymphoedema syndrome |
| **I97.8** | Other postprocedural disorders of circulatory system, not elsewhere classified |
| **I97.9** | Postprocedural disorder of circulatory system, unspecified |
| **J95.0** | Tracheostomy malfunction |
| **J95.1** | Acute pulmonary insufficiency following thoracic surgery |
| **J95.2** | Acute pulmonary insufficiency following nonthoracic surgery |
| **J95.3** | Chronic pulmonary insufficiency following surgery |
| **J95.4** | Mendelson syndrome |
| **J95.5** | Postprocedural subglottic stenosis |
| **J95.8** | Other postprocedural respiratory disorders |
| **J95.9** | Postprocedural respiratory disorder, unspecified |
| **K91.0** | Vomiting following gastrointestinal surgery |
| **K91.1** | Postgastric surgery syndromes |

| | |
|---|---|
| **K91.2** | Postsurgical malabsorption, not elsewhere classified |
| **K91.3** | Postoperative intestinal obstruction |
| **K91.4** | Colostomy and enterostomy malfunction |
| **K91.5** | Postcholecystectomy syndrome |
| **K91.8** | Other postprocedural disorders of digestive system, not elsewhere classified |
| **K91.9** | Postprocedural disorder of digestive system, unspecified |
| **L89.0** | Stage I decubitus ulcer and pressure area |
| **L89.1** | Stage II decubitus ulcer |
| **L89.2** | Stage III decubitus ulcer |
| **L89.3** | Stage IV decubitus ulcer |
| **L89.9** | Decubitus ulcer and pressure area, unspecified |
| **M96.0** | Pseudarthrosis after fusion or arthrodesis |
| **M96.1** | Postlaminectomy syndrome, not elsewhere classified |
| **M96.2** | Postradiation kyphosis |
| **M96.3** | Postlaminectomy kyphosis |
| **M96.4** | Postsurgical lordosis |
| **M96.5** | Postradiation scoliosis |
| **M96.6** | Fracture of bone following insertion of orthopaedic implant, joint prosthesis, or bone plate |
| **M96.8** | Other postprocedural musculoskeletal disorders |
| **M96.9** | Postprocedural musculoskeletal disorder, unspecified |
| **N99.0** | Postprocedural renal failure |
| **N99.1** | Postprocedural urethral stricture |
| **N99.2** | Postoperative adhesions of vagina |
| **N99.3** | Prolapse of vaginal vault after hysterectomy |
| **N99.4** | Postprocedural pelvic peritoneal adhesions |
| **N99.5** | Malfunction of external stoma of urinary tract |
| **N99.8** | Other postprocedural disorders of genitourinary system |
| **N99.9** | Postprocedural disorder of genitourinary system, unspecified |
| **T80.0** | Air embolism following infusion, transfusion and therapeutic injection |
| **T80.1** | Vascular complications following infusion, transfusion and therapeutic injection |
| **T80.2** | Infections following infusion, transfusion and therapeutic injection |
| **T80.3** | ABO incompatibility reaction |
| **T80.4** | Rh incompatibility reaction |
| **T80.5** | Anaphylactic shock due to serum |
| **T80.6** | Other serum reactions |
| **T80.8** | Other complications following infusion, transfusion and therapeutic injection |
| **T80.9** | Unspecified complication following infusion, transfusion and therapeutic injection |
| **T81.0** | Haemorrhage and haematoma complicating a procedure, not elsewhere classified |
| **T81.1** | Shock during or resulting from a procedure, not elsewhere classified |

| | |
|---|---|
| **T81.2** | Accidental puncture and laceration during a procedure, not elsewhere classified |
| **T81.3** | Disruption of operation wound, not elsewhere classified |
| **T81.4** | Infection following a procedure, not elsewhere classified |
| **T81.5** | Foreign body accidentally left in body cavity or operation wound following a procedure |
| **T81.6** | Acute reaction to foreign substance accidentally left during a procedure |
| **T81.7** | Vascular complications following a procedure, not elsewhere classified |
| **T81.8** | Other complications of procedures, not elsewhere classified |
| **T81.9** | Unspecified complication of procedure |
| **T82.0** | Mechanical complication of heart valve prosthesis |
| **T82.1** | Mechanical complication of cardiac electronic device |
| **T82.2** | Mechanical complication of coronary artery bypass and valve grafts |
| **T82.3** | Mechanical complication of other vascular grafts |
| **T82.4** | Mechanical complication of vascular dialysis catheter |
| **T82.5** | Mechanical complication of other cardiac and vascular devices and implants |
| **T82.6** | Infection and inflammatory reaction due to cardiac valve prosthesis |
| **T82.7** | Infection and inflammatory reaction due to other cardiac and vascular devices, implants and grafts |
| **T82.8** | Other specified complications of cardiac and vascular prosthetic devices, implants and grafts |
| **T82.9** | Unspecified complication of cardiac and vascular prosthetic device, implant and graft |
| **T83.0** | Mechanical complication of urinary (indwelling) catheter |
| **T83.1** | Mechanical complication of other urinary devices and implants |
| **T83.2** | Mechanical complication of graft of urinary organ |
| **T83.3** | Mechanical complication of intrauterine contraceptive device |
| **T83.4** | Mechanical complication of other prosthetic devices, implants and grafts in genital tract |
| **T83.5** | Infection and inflammatory reaction due to prosthetic device, implant and graft in urinary system |
| **T83.6** | Infection and inflammatory reaction due to prosthetic device, implant and graft in genital tract |
| **T83.8** | Other complications of genitourinary prosthetic devices, implants and grafts |
| **T83.9** | Unspecified complication of genitourinary prosthetic device, implant and graft |
| **T84.0** | Mechanical complication of internal joint prosthesis |
| **T84.1** | Mechanical complication of internal fixation device of bones of limb |
| **T84.2** | Mechanical complication of internal fixation device of other bones |
| **T84.3** | Mechanical complication of other bone devices, implants and grafts |
| **T84.4** | Mechanical complication of other internal orthopaedic devices, implants and grafts |
| **T84.5** | Infection and inflammatory reaction due to internal joint prosthesis |
| **T84.6** | Infection and inflammatory reaction due to internal fixation device [any site] |
| **T84.7** | Infection and inflammatory reaction due to other internal orthopaedic prosthetic devices, implants and grafts |
| **T84.8** | Other complications of internal orthopaedic prosthetic devices, implants and grafts |
| **T84.9** | Unspecified complication of internal orthopaedic prosthetic device, implant and graft |
| **T85.0** | Mechanical complication of ventricular intracranial (communicating) shunt |
| **T85.1** | Mechanical complication of implanted electronic stimulator of nervous system |

| | |
|---|---|
| **T85.2** | Mechanical complication of intraocular lens |
| **T85.3** | Mechanical complication of other ocular prosthetic devices, implants and grafts |
| **T85.4** | Mechanical complication of breast prosthesis and implant |
| **T85.5** | Mechanical complication of gastrointestinal prosthetic devices, implants and grafts |
| **T85.6** | Mechanical complication of other specified internal prosthetic devices, implants and grafts |
| **T85.7** | Infection and inflammatory reaction due to other internal prosthetic devices, implants and grafts |
| **T85.8** | Other complications of internal prosthetic devices, implants and grafts, not elsewhere classified |
| **T85.9** | Unspecified complication of internal prosthetic device, implant and graft |
| **T86.0** | Bone-marrow transplant rejection |
| **T86.1** | Kidney transplant failure and rejection |
| **T86.2** | Heart transplant failure and rejection |
| **T86.3** | Heart-lung transplant failure and rejection |
| **T86.4** | Liver transplant failure and rejection |
| **T86.8** | Failure and rejection of other transplanted organs and tissues |
| **T86.9** | Failure and rejection of unspecified transplanted organ and tissue |
| **T87.0** | Complications of reattached (part of) upper extremity |
| **T87.1** | Complications of reattached (part of) lower extremity |
| **T87.2** | Complications of other reattached body part |
| **T87.3** | Neuroma of amputation stump |
| **T87.4** | Infection of amputation stump |
| **T87.5** | Necrosis of amputation stump |
| **T87.6** | Other and unspecified complications of amputation stump |
| **T88.0** | Infection following immunization |
| **T88.1** | Other complications following immunization, not elsewhere classified |
| **T88.2** | Shock due to anaesthesia |
| **T88.3** | Malignant hyperthermia due to anaesthesia |
| **T88.4** | Failed or difficult intubation |
| **T88.5** | Other complications of anaesthesia |
| **T88.6** | Anaphylactic shock due to adverse effect of correct drug or medicament properly administered |
| **T88.7** | Unspecified adverse effect of drug or medicament |
| **T88.8** | Other specified complications of surgical and medical care, not elsewhere classified |
| **T88.9** | Complication of surgical and medical care, unspecified |
| **U69.0** | Elsewhere classified hospital-acquired pneumonia |
